# Optimal Testing Strategy for the Identification of COVID-19 Infections

**DOI:** 10.1101/2020.07.20.20157818

**Authors:** Michail Chatzimanolakis, Pascal Weber, George Arampatzis, Daniel Wälchli, Ivica Kičić, Petr Karnakov, Costas Papadimitriou, Petros Koumoutsakos

## Abstract

The systematic identification of infectious, yet unreported, individuals is critical for the containment of the COVID-19 pandemic. We present a strategy for identifying the location, timing and extent of testing that maximizes information gain for such infections. The optimal testing strategy relies on Bayesian experimental design and forecasting epidemic models that account for time dependent interventions. It is applicable at the onset and spreading of the epidemic and can forewarn for a possible recurrence of the disease after relaxation of interventions. We examine its application in Switzerland and show that it can provide timely and systematic guidance for the effective identification of infectious individuals with finite testing resources. The methodology and the open source code are readily adaptable to countries around the world.

*We present a strategy for the optimal allocation of testing resources in order to detect COVID-19 infections in a country’s population*.

## Introduction

The identification of unreported individuals infected by the SARS-CoV-2 virus is critical for the quantification and forecasting of the COVID-19 pandemic (*1–3*). Presently the spread of the disease is quantified by the reported numbers of infections, hospitalizations, recoveries and deaths. In turn, these quantities inform epidemiology models that provide short term forecasts for the spread of the epidemic and help quantify the role of possible interventions. The veracity of these forecasts depends on the discrepancy between the numbers of reported and unreported, yet infectious, individuals. The estimation of such unreported infections has been the subject of important testing campaigns (*4*). A challenge for these campaigns is that their estimates rely on testing symptomatic individuals or individuals that have been selected based on certain a priori selected criteria (hospital visits, airport arrivals, geographic vicinity to researchers, etc.) as well as on randomized tests of the population (*5*). There is broad recognition that efficient testing strategies are critical for the timely identification of infectious individuals and the optimal allocation of resources (*6–8*). However, targeted testing entails bias while randomized tests require access to a high percentage of the population with commensurate high costs.

Here we present an Optimal Testing Strategy (OpTS) that provides maximum information gain over the prior knowledge regarding infections. The method employs forecasts by epidemiological models (*9*) with parameters that are adjusted as data become available through a Bayesian inference framework. The forecasts are combined with Bayesian experimental design (*10–13*) to determine optimal testing parameters. The resulting OpTS is applicable in all stages of the pandemic, regardless of the availability of data.

We employ the *SEI*^*r*^*I*^*u*^*R* model (*14*) that predicts the number of susceptible (*S*), exposed (*E*), infectious reported (*I*^*r*^), unreported (*I*^*u*^), and removed (*R*) individuals from the population. The model amounts to a set of coupled ordinary differential equations with parameters inferred through a Bayesian framework. The *SEI*^*r*^*I*^*u*^*R* models the spread of a disease in a country’s population distributed in a number of communities that are interacting through mobility networks. Here we focus on Switzerland and consider its cantons as the respective communities. The model parameters are: the relative transmission rate (*µ*), the virus latency period (*Z*), the infectious period (*D*) and the reporting rate (*α*). Moreover the transmission rate (*β*) and the mobility factor (*θ*) are considered to be time dependent in order to account for government interventions.

Bayesian inference is used to quantify and propagate uncertainties (*15*) in the model parameters. At the onset of the epidemic, the uncertainty is quantified solely through prior probability distributions. As daily infections data become available, the model is updated by estimating the posterior distribution of its parameters using Bayesian inference. These distributions are used to propagate uncertainties in the model forecasts, informing decision makers regarding the containment of the disease (*9*). We note that the uncertainty in the model parameters and initial conditions has a profound effect on predictions of the disease dynamics and associated uncertainty bounds (*16*). Large uncertainty around the most probable parameter values may result in large uncertainty bounds for critical quantities of interest, thus hindering the decision process for identifying effective interventions.

The OpTS aims to acquire the most informative data, to reduce model uncertainties, with limited test resources. Minimizing the uncertainty of the model parameters leads to more reliable predictions for quantities such as the reproduction number (*17*). Moreover, it results in state variables with smaller uncertainty bounds, thus reducing risks associated with the decision making process including timing, extent of interventions and probability of exceeding hospital capacity. Here the OpTS provides optimal estimates for times and selected communities to estimate the number of unreported infectious individuals in a country’s (Switzerland) population. We measure the information gain from these tests using a utility function (*18–21*) based on the Kullback-Leibler divergence between the inferred posterior distribution and the current prior distribution of the model parameters. We remark that the prior can be formulated using the posterior distribution estimated from daily data of the infectious reported individuals up to the current date (see Materials and Methods). Hence, in any stage of the epidemic we can estimate the optimal time and location/community where testing has to be carried out to maximize the expected information gain regarding infections in a population.

We show the effectiveness of the proposed OpTS over non-specific, randomized testing of sub-populations. Furthermore, we demonstrate the capabilities of the method throughout the COVID-19 pandemic, from the onset of the epidemic to monitoring the effectiveness of interventions and forewarning for a possible second outbreak. Our results are based on the spread of the coronavirus disease in the cantons of Switzerland and an extended *SEI*^*r*^*I*^*u*^*R* epidemiological model that accounts for time dependent mitigation strategies (*22*). However, the testing method is readily applicable to other countries and can readily accommodate different epidemiological models.

### Optimal Testing during the COVID-19 pandemic

We present OpTS for three different stages of the epidemic: (i) starting phase (blue), (ii) containment after enforcement of interventions (red) and (iii) relaxing of interventions and monitoring for a possible second outbreak (green) (Fig.1). The strategy relies on Bayesian experimental design and can operate when no data are available (as in the start of the epidemic) as well as when data have been accumulated, as in the last two stages of the epidemic. Testing campaigns rely on acquiring randomized samples from a population. The collected data, together with epidemiological models, help determine quantities of interest, such as the basic reproduction number of the disease (*17*). By suitably adapting the testing campaign, the data can help reduce the model uncertainty, thus enabling accurate forecasts and improved estimates regarding the severity of an epidemic.

A testing campaign consists of a set (***s***) of randomized tests *s*_*i*_ = (*k*_*i*_, *t*_*i*_), *i* = 1, … *M*_*y*_ performed in location *k*_*i*_ and on day *t*_*i*_. The expected information gain of a particular strategy for selecting the testing locations/times ***s*** is quantified by using a utility function *Û* (***s***) (*18*). The maximum of this function corresponds to an optimal strategy that yields the most information about the quantities of interest. A key challenge for OpTS is their computational cost (*12*). Here, the OpTS is rendered computationally efficient through a sequential optimization algorithm (*23*) that adds tests iteratively.

The OpTS relies the characterization and prediction of the spread of the disease based on forecasts by suitable epidemiological models (*22*). In turn, these forecasts rely on prior information and their predictions are further adjusted as data become available in a Bayesian inference framework (*9*). Here, the set of Ordinary Differential Equations (ODEs) describing the *SEI*^*r*^*I*^*u*^*R* model (*14*) are integrated to produce the model output. The uncertainty of the model output and its discrepancy from the available data is quantified through a parametrized error model. The stochastic model and its quantified uncertainties are then used to identify the OpTS. The reader is referred to the Supplementary Materials of this paper for the definition of the utility function, the optimization algorithm as well as for the epidemiological and error models.

### Beginning of the epidemic - Optimal testing without data

At the start of an epidemic, there are no data and we assume no other prior information regarding the spread of the pathogen in a country. The initial conditions for the number of unreported infections 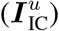 were selected with non-zero values for the cantons of Aargau, Bern, Basel-Landschaft, Basel-Stadt, Fribourg, Geneva, Grisons, St.Gallen, Ticino, Vaud, Valais and Zurich based on their population and their large number of interconnections. Due to the lack of any prior information and relevant data, all the parameters are assumed to follow uniform prior distributions (see table S5, for details).

In Switzerland the first infectious person was reported on February 25^th^ in the canton of Ticino 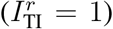 with no initial reported infections in all other cantons. The initial number of exposed individuals is set proportional to the number of unreported infections 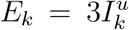 in accordance with the value of *R*_0_ ≈ 3 reported in (*24*) in the initial stage of the disease. The rest of the population is assumed to be susceptible. The methodology involves parameters of interest (***ϑ*** = (*β, µ, α, Z, D, θ, c*)) and nuisance parameters 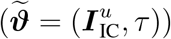 that the testing strategy does not aim to determine (see Supplementary Material for definitions).

The estimated expected utility functions 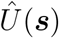 for up to four measurements in the cantons of Switzerland for a time horizon of 8 days is shown in Figure 2. Higher values for expected utility are estimated in cantons with larger population reflecting the larger relative uncertainty for cantons with only few reported cases. This implies that small cantons with little mobility rates are less preferred for performing tests since their contribution to the information gain is not significant. The Bayesian analysis enables the inference of the particular cantons and days for which tests should be performed in order to maximize the information gain. Accordingly, the most informative measurement should have been taken in Zurich on March the 2^nd^. Following the sequential optimization strategy, the optimal location and time for the second measurement is determined to be canton of Vaud on the 27^th^ of February. As expected, the information that is gained from tests in the canton of Vaud is less than the information gained from the canton of Zurich. The information that would have been gained by testing the next two preferred cantons of Vaud and Basel-Landschaft on March the 3^rd^ and February the 28^th^ respectively, is progressively reduced to a small level that, given the testing costs, does not justify carrying out tests in more than 4 cantons. The values of the optimal times are listed in table S1 in the Supplementary Materials.

**Figure 2:**
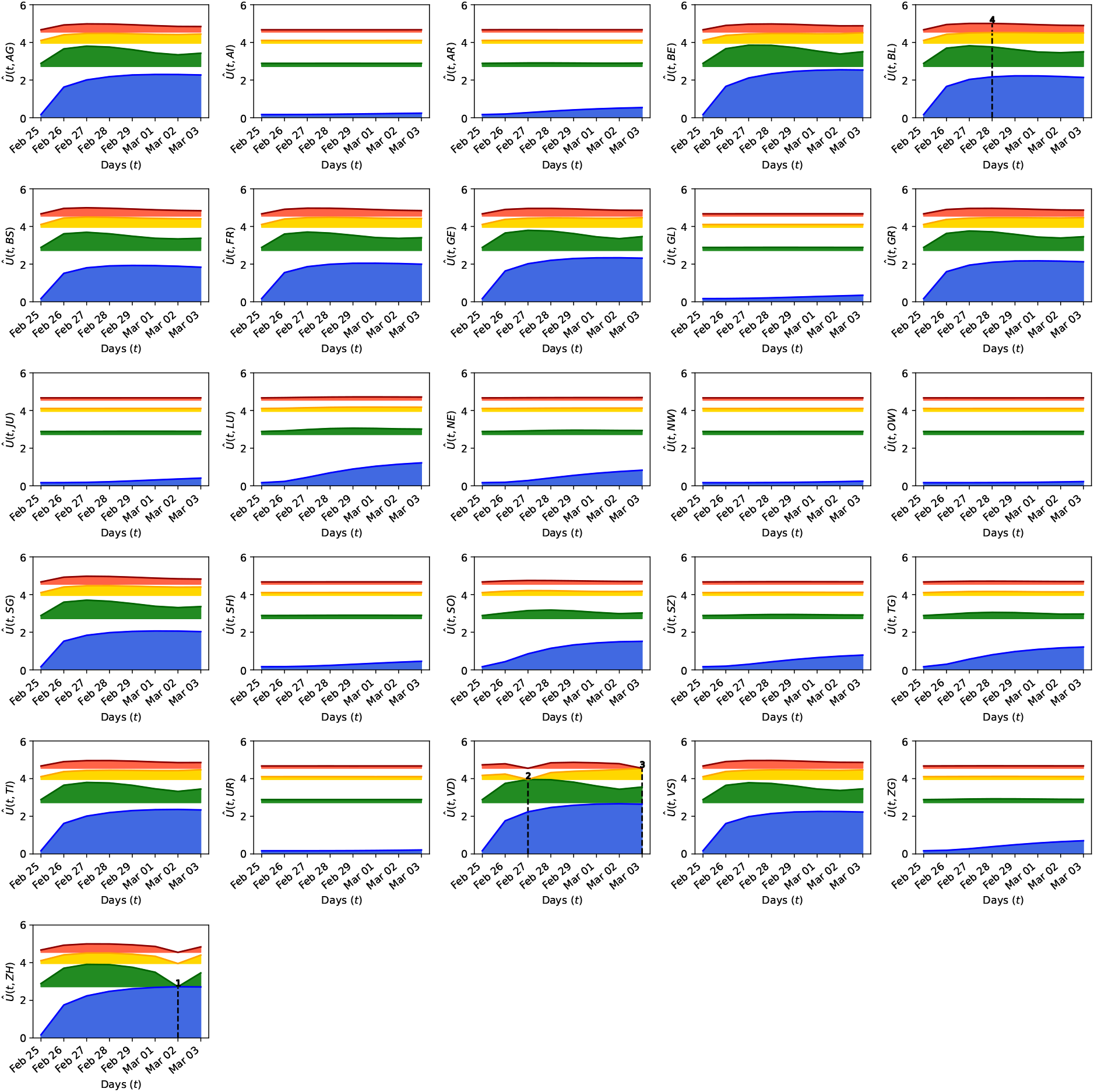
Expected information gain during start of epidemic. The blue curve corresponds to the utility of taking one measurement. The green curve is the utility when a second measurement is added, provided the location and time of the first measurement correspond to the maximum of the blue curve (found in the canton of Zurich, on March 2^nd^). Similarly, the yellow and red curves show the utilities for a third and fourth measurements, when the locations and time of the previous measurements are fixed to their optimal values. The fixed dates and location of each measurement are plotted with black dashed lines. The shaded areas indicate the difference to the expected information gain of the previous measurement, which becomes thinner as additional measurements do not yield a further significant information gain.

The results indicate that the proposed OpTS selects certain populous and well interconnected cantons at specific times to acquire the most information for estimating the model parameters.

### Exponential spreading and optimal testing strategy during non-pharmaceutical interventions

When the spreading of the coronavirus entered an exponential growth stage, several governments decided to take non-pharmaceutical interventions such as requesting social distancing, closing schools and restaurants, or even ordering a complete lockdown in order to contain the epidemic. Here, the goal of the OpTS is to obtain measurements that they help to better assess the effectiveness of these interventions.

In this case, priors for the model parameters are informed using data from the spread of the COVID-19. The daily reported infections in Switzerland (*25*) from the 25^th^ of February up to the 17^th^ of March 2020 are used to update the uniform prior distributions, specified in the previous phase, by using Bayesian inference (see figure S1 in Supplementary Materials). In the *SEI*^*r*^*I*^*u*^*R* model we model the non-pharmaceutical interventions with a time-dependent infection rate *β* and the mobility factor *θ*. These parameters are also calibrated by the data and provide an estimate on the timing and effectiveness of the interventions (*16*).

Figure 3 shows the maximum values of the information gain for each measurement. For cantons with a small population and low connectivity to other cantons a low information gain is found. The opposite can be observed for cantons with large population and strong connections to other cantons. The values for the maximum utility in time for the measurements are listed in Table S2.

**Figure 3:**
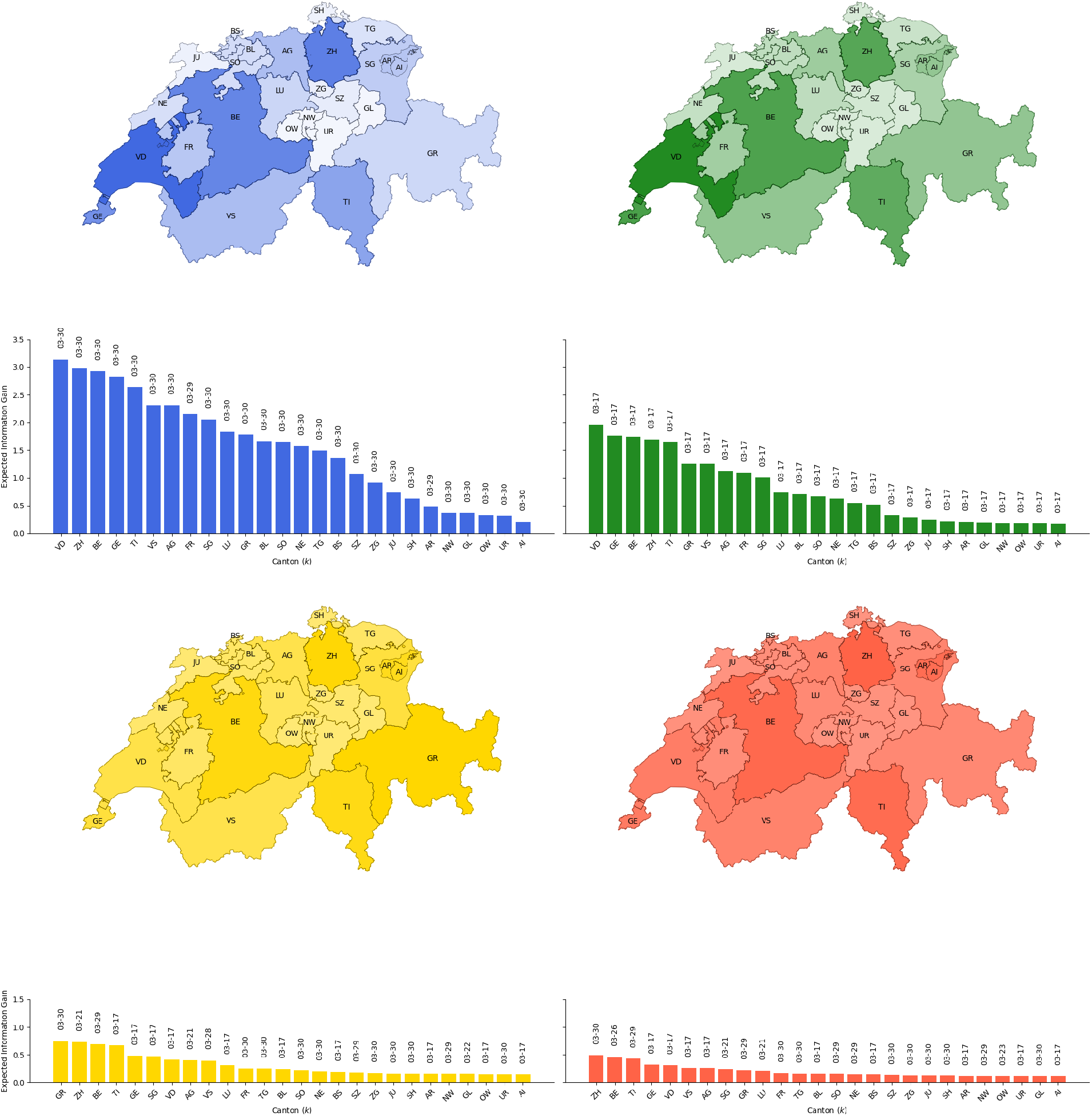
Optimal testing strategy for effect of non-pharmaceutical interventions. The maximum gain of information is plotted on the map of Switzerland. Here blue corresponds to taking one measurement, green to adding a second, yellow to a third and red to a fourth. Below the map we plot the magnitude of the expected information gain of each measurement, along with the optimal measurement dates per canton.

If only a single canton is to be selected, tests in the canton of Vaud carried out on the 30^th^ of March are to be preferred over tests in either of the cantons of Zurich, Bern or Geneva (blue in figure 3). If two tests can be afforded, the OpTS is to carry them out in the same canton (Vaud) on the 17^th^ and on the 30^th^ March (blue and green in figure 3). Note that the canton of Zurich, ranked as the next preferred canton for a single test (blue in figure 3), is not selected by the methodology since part of the information that would be gained from testing is already contained in tests performed in Vaud. The optimal location and time for a third test is the canton of Grisons on the 30^th^ of March (yellow in figure 3). The canton of Zurich is proposed as the fourth location to be tested on the 30^th^ of March as well. However, the information gain from the fourth test in the canton of Zurich is approximately 10% of the total information gained from the tests carried optimally in the first three cantons.

The results suggest that tests in two locations/times provide significant information regarding assessing the effectiveness of interventions. Further tests on more locations/times do not add substantial information. It is evident that a trade-off between the required information gain and cost of testing are decisive for the number of necessary tests.

### Optimal monitoring for a second outbreak

After the relaxation of measures that assisted in mitigating the initial spread of the disease, it is critical to monitor the population for a possible second outbreak (*5*). The proposed OpTS supports such monitoring with randomized tests of the population based on data up to and after the release of the measures.

First, Bayesian inference is performed with data available up to June the 6^th^, allowing for an update of the uniform priors. This date is in accordance to the first stage of major release of measures in Switzerland (*26*). The effects of interventions are modeled by a parametrized time-dependent infection rate and mobility factor (see Supplementary Materials for a detailed description). The inferred probability distributions of these additional parameters are taken into account as the OpTS maximizes the information gain.

Subsequently, data up to July the 9^th^ are included, repeating the Bayesian inference and estimating the OpTS (marginal distributions and predictions shown in Figures S3 and S4). The inference indicates that the relaxation of measures correlates with an increase in the number of reported infections as observed in Figure 4. The information gain for each canton indicates the most informative tests should be performed a week after performing the inference. The provided information could then assist in estimating the severity of a second outbreak as indicated by the maximum of the utility in time (Tables S3 and S4).

**Figure 4:**
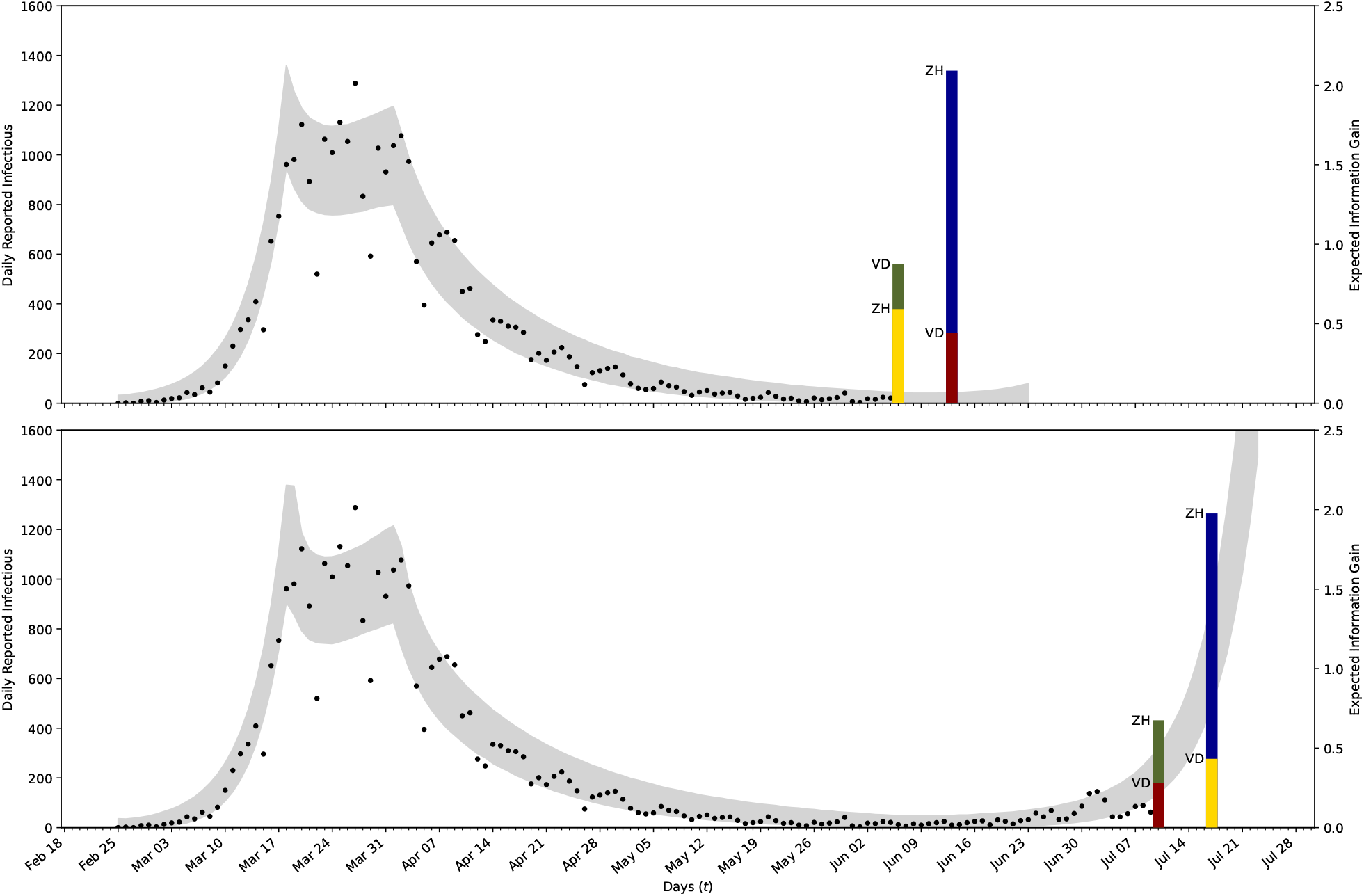
Optimal testing strategy to monitor a second outbreak. We use Bayesian inference to determine the parameters of the first infection wave using the data (black dots) of the daily new reported infections up to the 6^th^ of June (upper plot) and to the 9^th^ of July (lower plot). The 99% confidence intervals are plotted in gray. The proposed testing strategy is plotted with vertical bars at the found optimal days. Here blue indicated the utilities for the first measurement. The green bars correspond to the gain in utility when adding a second measurement assuming the first was chosen in the optimal location, where the yellow and red correspond to adding a third and fourth measurement.

Given that tests should be carried out in four locations and times, the methodology promotes optimal tests for two different times, within a week, in the cantons of Zurich and Vaud. First, tests should be performed in Zurich, providing high information gain for both considered cases. The next two tests are to be performed in Zurich and Vaud, with a rank that depends on the considered case, while the fourth test should be performed in Vaud. We find that the information gain from the last test is approximately 10% of the cumulative information gain from the first three tests. The number of tests can be then selected according to the available resources.

### Effectiveness of Optimal Testing

We demonstrate the importance of following an OpTS by comparing it with a non-specific testing campaign. We first re-examine the situation at the start of an epidemic and assume that the available resources allow for two randomized tests. Tests are simulated by evaluating the epidemiological model with the maximum a-posteriori estimate (MPE) of the parameters obtained from the inference in phase II (exponential growth) of the epidemic. We used data of the first 21 days of the infection spread in Switzerland (*25*) (February 25^th^ to March 17^th^). After evaluating the model, artificial measurements are obtained by adding a stochastic error term.

For the optimal strategy, data are collected by consulting figure 2. Thus, the two randomized tests are performed in the cantons of Zurich and Vaud, on the 2^nd^ of March and the 27^th^ of February respectively. For a non-specific strategy, the cantons of Ticino and Bern were selected, on the 28^th^ of February. These artificial data, obtained for the two strategies, are added to the real data of the daily reported cases from the first 8 days after the outbreak in Ticino. For the expanded data-set 𝒟 the posterior distributions 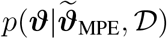 are found by sampling the model parameters using nested sampling (*17*). Note that the value of *c* is also inferred. For simplicity, the value of the correlation time *τ* is assumed to be known as this does not influence the results as long as the tests are carried out in different cantons.

The resulting one- and two-dimensional marginalized posterior distributions for both strategies are shown in figure 5. We note that the dispersion coefficient *r* (defined in the Supplementary Materials) in the error model for the real data (the reported infections) and the correlation parameter are almost the same for both strategies. However the model parameters show significant differences even when only two new data-points are added to a set of 208 data-points.

**Figure 5:**
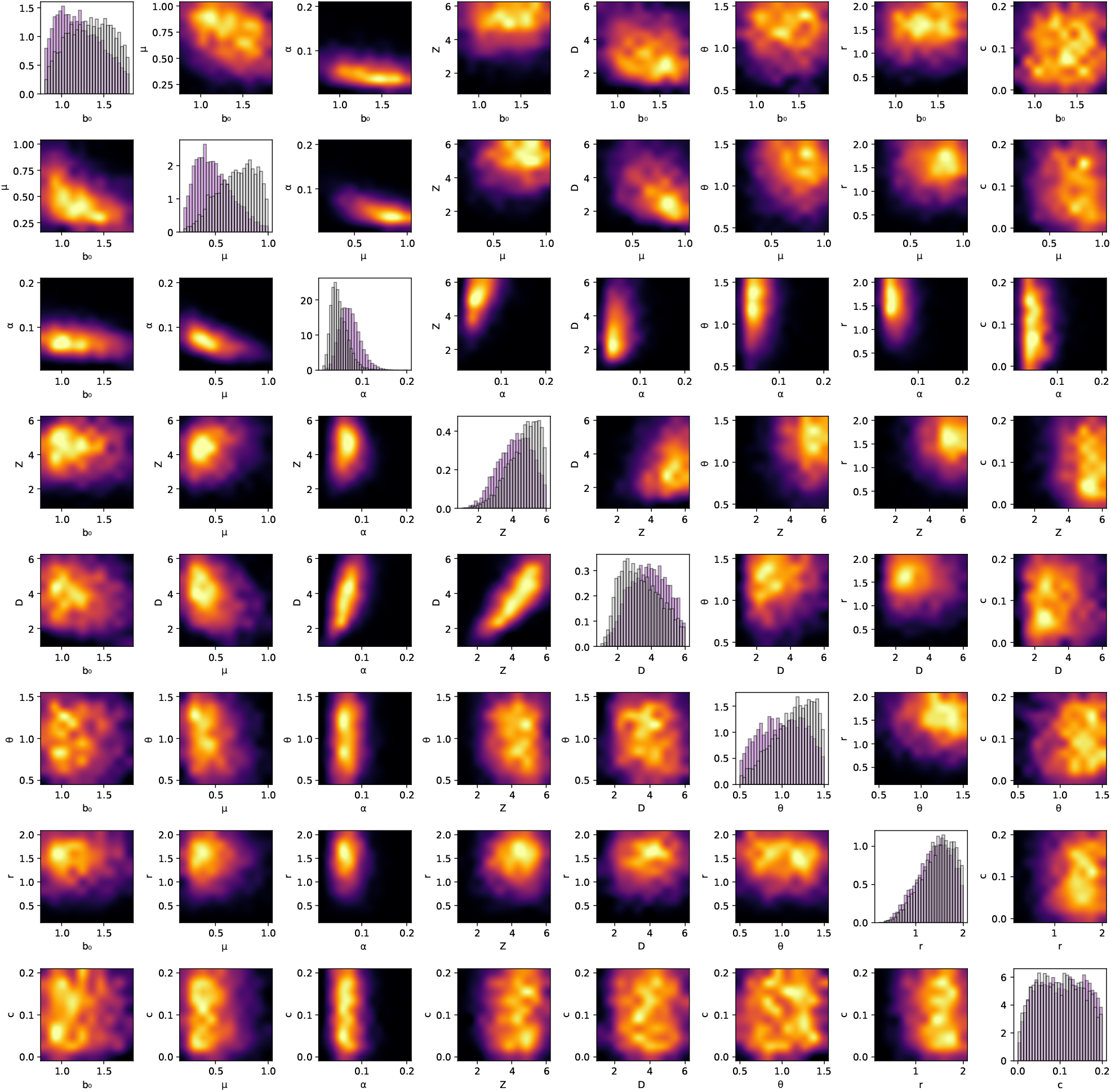
Marginal posterior distributions for two strategies. The diagonal shows the histogram for the marginal distribution for every parameter. Purple indicates posterior for the the measurement following the optimal testing strategy, gray the one for the non-specific strategy. The lower half and upper half show the samples of the joint distribution of two parameters for the optimal and the non-specific strategy respectively. Here black indicates low density and yellow high density.

The posterior distributions of the parameters of interest are propagated through the epidemiology model to provide the uncertainties in the number of unreported infectious individuals. In figure 6 the model output for the total number of unreported infections is plotted together with a 99% confidence interval along with the true value of the unreported cases obtained by using the selected parameters. The predictions from the OpTS have a much higher certainty with a confidence interval that is up to four times narrower than the one from a non-specific strategy. Further comparisons, demonstrating the value of the OpTS, include model predictions with higher certainty, as indicated by confidence intervals that are narrower than the ones obtained from from a non-specific strategy (figures S4 and S5, see Supplementary Material). Narrower uncertainty bounds provide higher confidence for decisions related to possible interventions to contain the epidemic.

**Figure 6:**
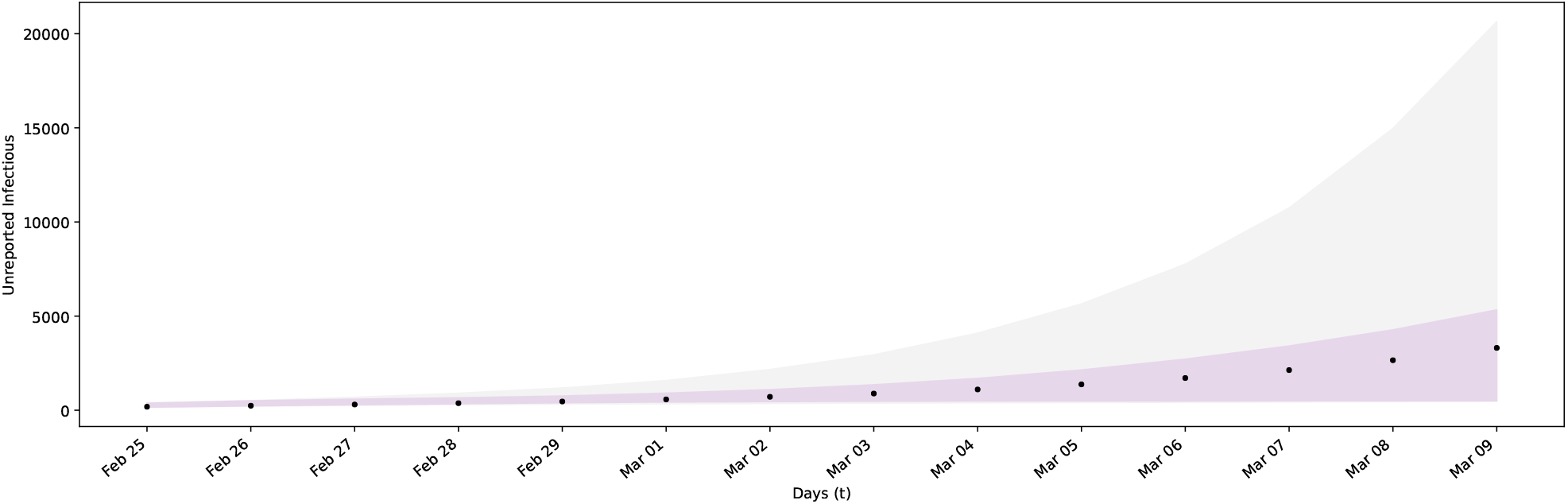
Prediction uncertainty for different testing strategies. The black dots show the actual unreported infectious for an artificial spread in Switzerland. The error bounds show the 99% confidence intervals of the model output for samples of the parameters with data obtained by optimal (purple) and non-specific testing (gray).

## Discussion

We introduced a systematic approach to identify optimal times and locations for randomized tests in order to quantify infectious individuals of a country’s population during the COVID-19 epidemic. The proposed OpTS exploits prior information and available data to maximize the expected information gain in parameters of interest and to minimize uncertainties in the forecasts of epidemiological models. In turn, improved forecasts for the virus spread provide rational guidelines for optimal allocation of finite testing resources.

The proposed method is demonstrated by focusing on the outbreak of the epidemic in Switzerland. The methodology relies on Bayesian experimental design using prior information and available data of reported infections along with forecasts from the *SEI*^*r*^*I*^*u*^*R* model. We compute the optimal testing strategy for three phases of the epidemic. First, we quantify the spread of the disease after the first reported infection. At this stage the method identifies the most crucial dates and locations for randomized tests in the country’s population. The deployment of OpTS at this phase would have allowed authorities to perform randomized testing in a period of high uncertainty, well in advance of the disease outbreak. During the period of interventions the proposed strategy would help quantify their effectiveness assisting decision making for further interventions or retraction of measures that may be harmful to the economy. Finally, the OpTS can assist monitoring for a recurrence of the disease after preventive measures have been relaxed and help guide further planing of interventions.

We remark that the proposed OpTS does not depend on a particular type of data/model or to the country of Switzerland. The open source code is modular, scalable and readily adaptable to different scenarios for the epidemic and countries around the world. We believe that the present work can be a valuable tool for decision makers to allocate resources efficiently for testing the population, providing a reliable quantification of the spread of the disease and designing effective interventions.

## Data Availability

All data is available in the manuscript or the Supplementary Materials.

## Acknowledgments

We acknowledge discussions with Fabian Wermelinger, Lucas Amoudruz, Martin Boden (ETHZ). Sergio Martin (ETHZ) provided technical assistance with the software.

## Authors contributions

Conceptualization: C.P., P.Ko.; Data curation: M.C., P.W.; Formal Analysis: M.C., P.W., G.A., D.W., C.P.; Funding acquisition: P.Ko.; Investigation: M.C., P.W., C.P., G.A., P.Ko. Methodology: P.W., M.C., C.P., G.A., P.Ko.; Project administration: P.Ko.; Resources: P.Ko.; Software: P.W., M.C., D.W., I.K., P.Ka.; Supervision: C.P., P.Ko., G.A.; Validation: M.C., P.W.; Visualization: P.W., M.C., P.Ka; Writing – original draft: P.W., M.C., C.P., P.Ko.; Writing – review & editing: P.W., M.C., C.P., P.Ko., G.A., D.W., I.K., P.Ka.

## Data and materials availability

All data is available in the manuscript or the Supplementary Materials.

## Supplementary Materials for

### Materials and Methods

The optimal time (day) and location (canton) for testing a population to detect infectious individuals is determined via Bayesian optimal experimental design (*12, 13, 27*). This optimal testing strategy (OpTS) relies on combining Bayesian inference and utility theory with forecasting models of the epidemic. We remark that the OpTS does not depend on a particular epidemiological model or type of data. The methodology is applicable at all stages of the epidemic (inception to re-occurrence). It can operate without data at the early stages of the pandemic and takes advantage of data available at later stages of the pandemic. The methodology is rendered computationally efficient using a sequential optimization algorithm (*23*).

### Bayesian Inference from randomized testing

We consider a testing campaign including a set (***s***) of randomized tests *s*_*i*_ = (*k*_*i*_, *t*_*i*_), *i* = 1, … *M*_*y*_ performed in location *k*_*i*_ and on day *t*_*i*_. These tests measure a quantity of interest (QoI), that is denoted by 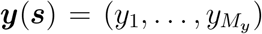. Here, *y*_*i*_ is the number of unreported infectious individuals, measured through test *s*_*i*_. The QoI can be predicted by a model 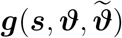 (here the *SEI*^*r*^*I*^*u*^*R* epidemiological model) that depends on parameters of interest ***ϑ*** ∈ ℝ ^*N*^ and nuisance parameters ***ϑ*** 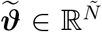. The distinction between model and nuisance parameters is discussed in later sections. We note that both sets of parameters are uncertain and the proposed method aims to reduce the uncertainty only in the parameters of interest.

A stochastic error term ***ε***(***s***) links the model prediction with the QoI :

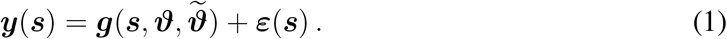

The error ***ε***(***s***) is assumed to follow a zero-mean multivariate normal distribution 𝒩 (0, Σ) with covariance matrix 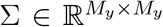. The elements of the covariance matrix (Σ_*s,s′*_) correspond to measurements taken at *s* = (*k, t*) and *s ′* = (*k* ′, *t* ′) and are given by

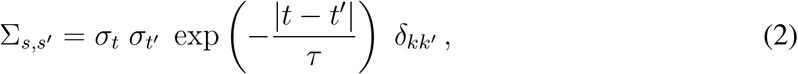

where *δ*_*kk ′*_ is the Kronecker delta, which is 1 for *k* = *k*′ and 0 otherwise. The correlation time *τ* ∈ [0.5, 3.5] is considered a nuisance parameter. These assumptions about the covariance imply that measurements in different locations are not correlated, while those in the same location have an exponentially decaying temporal correlation. The latter avoids clustering of measurements in small time intervals (*28, 29*). The factor *σ*_*t*_ ∈ ℝ is assumed proportional to the expectation of the QoI, taken over all possible test locations and over the range of model and nuisance parameters

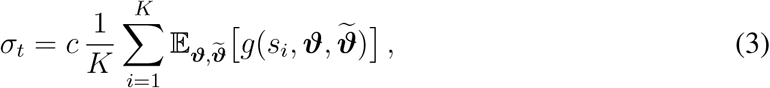

where *s*_*i*_ = (*i, t*). The parameter *c* ∈ [0, 0.25] is considered a model parameter. The expectation 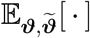 is taken with respect to all parameters ***ϑ*** and 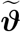 that follow the prior probability distribution with density 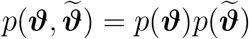.

Under these assumptions, the conditional probability of ***y*** on 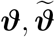 and ***s*** is given by

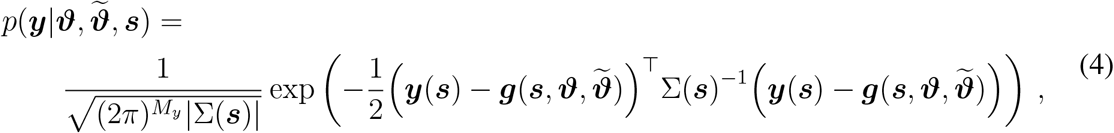

where |Σ(***s***)| is the determinant of the covariance matrix.

### Expected Information Gain

The most informative measurements ***y*** provide the least uncertainty in the estimates of the model parameters ***ϑ***. Starting with a user-postulated prior distribution *p*(***ϑ***), Bayesian learning is used to update the uncertainties in the model parameters leading to a posterior distribution 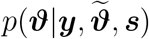, based on the information contained in the test data ***y***. The Kullback–Leibler (KL) divergence between the posterior 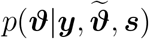 and the prior distributions *p*(***ϑ***) of the model parameters measures the distance between the two distributions.

Informative data produce posterior distributions that differ from the prior; greater differences lead to higher information gain. Therefore, the most informative data ***y*** correspond to the testing strategy (measurement locations and times) with the highest information gain (*18, 19*).

The OpTS is identified by maximizing a utility function (*12, 20*). One choice is the KL divergence 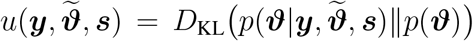 quantifying the information gain from the data (*12*). However, since data are not available in the experimental design phase, the utility function is selected here to be the expected KL divergence 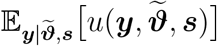 over all data generated by the model prediction error equation (1). Also, to account for the uncertainty in nuisance parameters 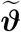, encoded in the prior distribution 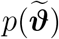, the expectation is also taken with respect to 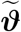, which results in the utility function (*20*)

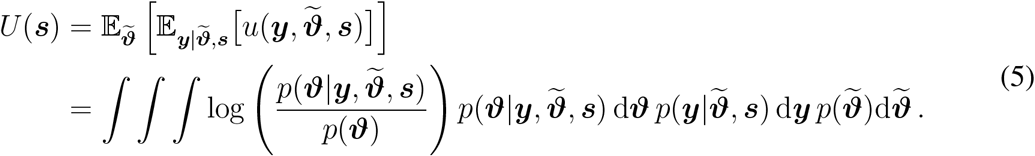

By using Bayes’ theorem

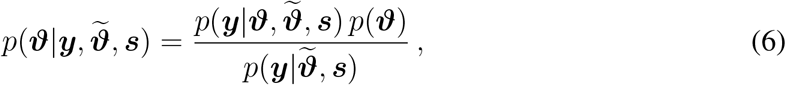

the utility function can be simplified to

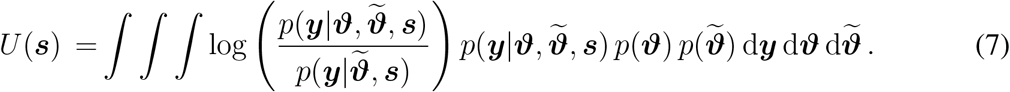

Note that the expected utility only depends on the locations and times of the measurements via ***s***. The term 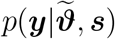 is the model evidence given by

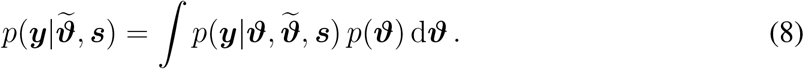

The choice of the prior distribution *p*(***ϑ***) for the parameters allows to incorporate prior knowledge from epidemiology. If no information is available from data, a case encountered in the beginning of the infection, a uniform prior distribution can be assumed. Table S5 summarizes our choice of prior distributions for all the involved uncertain quantities. If data ***d*** of the daily number of reported infectious individuals is available, Bayesian inference can be used to inform the prior distribution, as described later on. In this case, the prior *p*(***ϑ***) in equation (7) is replaced by the distribution *p*(***ϑ***|***d***) informed from the data ***d***.

In the present work, the assumed nuisance parameters are the correlation time *τ* and the initial condition of the unreported infections in the cantons of Aargau, Bern, Basel-Landschaft, Basel-Stadt, Fribourg, Geneva, Grisons, St.Gallen, Ticino, Vaud, Valais and Zurich 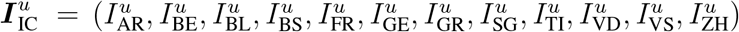, with prior distributions 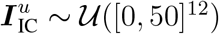 and *τ* ∼ U([0.5, 3.5]).

### Epidemiological Model

Here we employ the *SEI*^*r*^*I*^*u*^*R* epidemiological model (*14*) to forecast the dynamics of the coronavirus outbreak in Switzerland

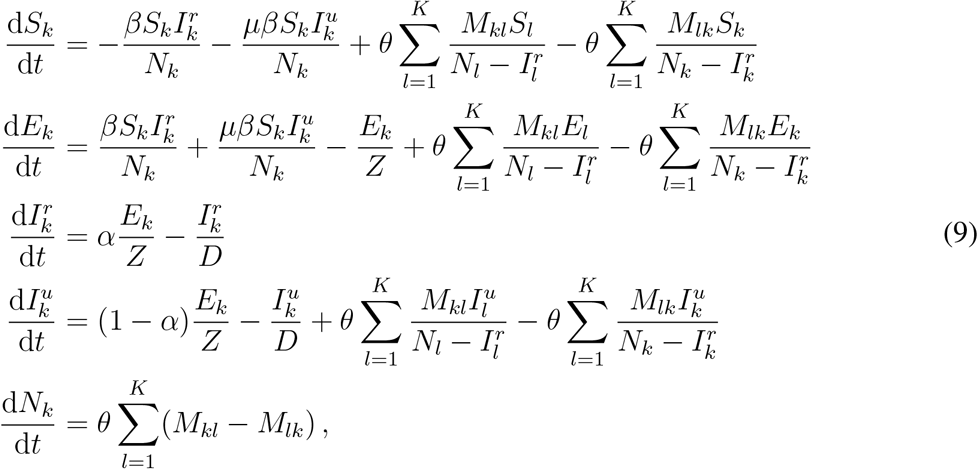

where 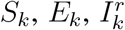 and 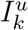 denote the number of individuals in canton *k* = {1, …, *K*} that are susceptible, exposed, reported infectious and unreported infectious, respectively. We denote by *K* the number of cantons (26 in Switzerland), by *N*_*k*_ the total population of the canton *k*, while the population mobility between cantons *k* and *l* is denoted by *M*_*kl*_ with values obtained from the Swiss Federal Statistical Office (*30*). The model parameters are the transmission rate (*β*), the relative transmission rate between reported and unreported infectious individuals (*µ*), the virus latency period (*Z*), the infectious period (*D*), the reporting rate (*α*) and the mobility factor (*θ*).

We employ different time-dependent expressions for the transmission rate and the mobility factor for each stage of the epidemic. Constants are chosen for the start of an epidemic while in the cases of monitoring of interventions, the following expressions are used:

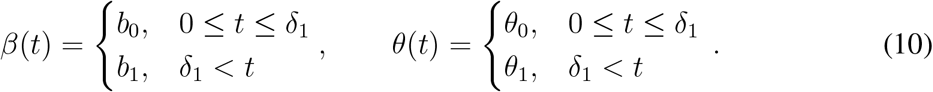

where *b*_0_, *b*_1_, *θ*_0_ and *θ*_1_ are the infection rates and mobility factors before and after the intervention. Time *t* = 0 corresponds to the 25^th^ of February 2020, and *δ*_1_ = 21 to the 17^th^ of March 2020, when the lockdown was announced in Switzerland (*26*). Finally, for the third case (monitoring of a second outbreak) we assume that

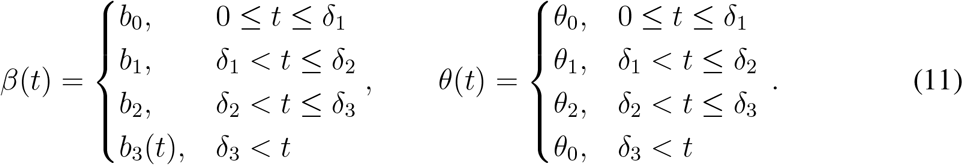

As in equation (10), *b*_0_ is the infection rate before the intervention while *b*_1_ = *c*_1_ *b*_0_ and *b*_2_ = *c*_2_ *b*_0_ with *c*_1_, *c*_2_ ∈ [0, 1] are the infection rates after the two interventions. Similarly, *θ*_0_ is the mobility factor before any interventions took place, while *θ*_1_ = *c*_3_ *θ*_0_ and *θ*_2_ = *c*_4_ *θ*_0_ with *c*_3_, *c*_4_ ∈ [0, 1] are the mobility factors after the two interventions. Moreover, *δ*_1_ and *δ*_2_ correspond to the days of the interventions. The day when the measures are loosened is denoted by *δ*_3_. After that day, the infection rate is gradually increasing

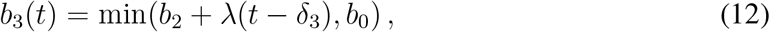

with *λ* ∈ [0, 0.03], while the mobility factor regains its initial value of *θ*_0_.

### Estimation of the Expected Information Gain

The calculation of the expected utility from equation (7) is performed with Monte-Carlo integration. Samples from the prior distribution are denoted by 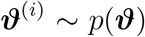 and by 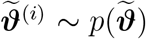, while samples on the measurement space are denoted by 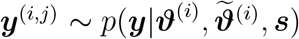, where *i* ∈ {1, …, *N*_***ϑ***_} and *j* ∈ {1, …, *N*_*y*_}. With these samples, an estimate of the expected utility is computed as

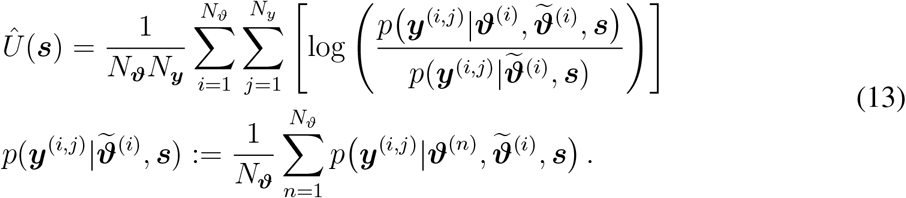

In our implementation the samples ***ϑ***^(*i*)^ and 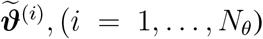, (*i* = 1, …, *N*_*θ*_), remain the same for different values of ***s***. Thus, the model evaluations 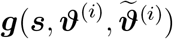 are only carried out once and are stored and used in the iteration process involved in the optimization. This allows to separate the computational cost of the model evaluation from the cost of computing the utility, which scales as 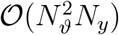.

### Optimal Location and Time of Testing

We define the optimal test times and locations as

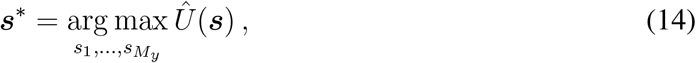

where 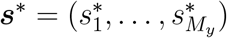 with 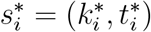 denote the locations 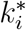 and times 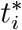 for the optimal measurements with *i* ∈ {1, …, *M*_*y*_}. For a grid search, the associated computational cost is 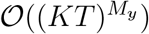 and thus grows exponentially with the number of tests. This curse of dimensionality is avoided by using a sequential optimization method (*23, 31, 32*) to approximate the global optimum by iteratively solving

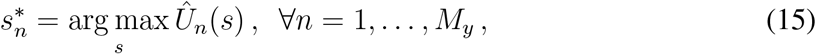

where *s* = (*k, t*) is the location and time to be estimated sequentially starting with *n* = 1 and

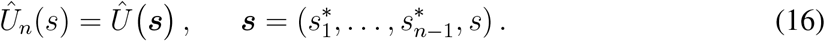

Following this, we define the expected information gain for testing sequence *n* as

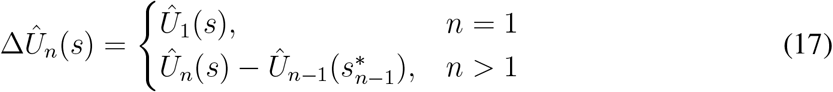

### Quantification of Uncertainty

A data informed prior *p*(***ϑ***|***d***) of the model parameters ***ϑ*** can be computed from available data 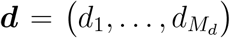, collected at *M*_*d*_ locations and days. Here, available data ***d*** refer to the daily number of reported infectious individuals and they are contrasted from the data ***y*** of the number of unreported infectious individuals. The latter are obtained from testing strategies at selected populations using optimal experimental design. The data is mapped via a distinct model output 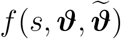 through the following error model

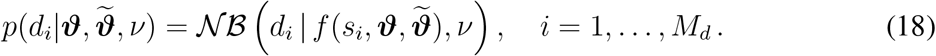

where 𝒩 ℬ is the negative binomial distribution with mean *f* and dispersion *ν*. Also, *s*_*i*_ = (*k*_*i*_, *t*_*i*_) is the location and time the data *d*_*i*_ was collected. The choice of a different error model, compared to equation (1), is based on the assumption that the data are independent and identically distributed. Such an assumption would not be acceptable in the measurement model in equation (1), as it may result in uncorrelated measurements that can become clustered in small time intervals (*28, 29*).

The data ***d*** = *d*_1_, …, *d*_*M*_*d* are the daily number of reported infections per canton in Switzerland (*25*) which corresponds to the following model quantity

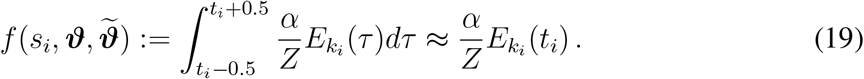

The posterior distribution that will be used subsequently as a data informed prior is obtained using Bayes’ theorem

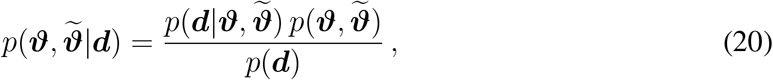

and is sampled with a nested sampling algorithm (*17*). Note the difference to equation (6) and the optimal testing methodology, where we are interested to reduce the uncertainty in 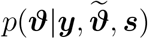, which excludes the nuisance parameters 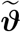. For the dispersion parameter in equation (18), it is assumed that 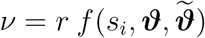. The coefficient *r* is unknown and included in the parameter set, where *r* ∼ 𝒰 ([0, 2]).

The three inferences performed are summarized in table S5, which shows the involved model parameters in each case. The histograms for the found samples are shown in figures S1, S2, and S3.

We remark that, using the present methodology, the inferred date for the beginning of the intervention is *δ*_1_ = 22.5, which is the 18^th^ of March 2020, corresponding well with the 17^th^ of March 2020 on which the lockdown was introduced in Switzerland (*26*). Moreover, we infer a significant reduction in the mobility factor, which indicates that traffic between cantons was also minimized. For the inference III we plot the fit using the inferred parameters in figure S4. The daily reported cases per canton are shown, together with the data used for the inference.

**Figure 1:**
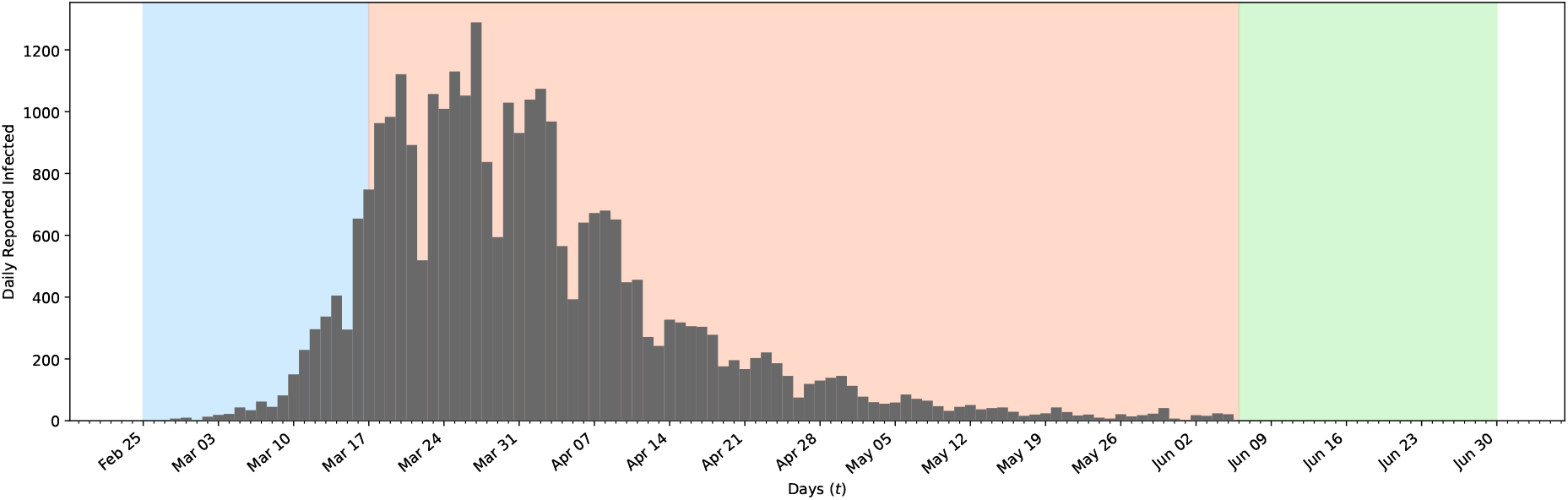
Testing scenarios for the COVID19 outbreak in Switzerland. Daily reported Coronavirus cases in Switzerland are plotted as gray bars. The period before (blue), during (red) and after (green) imposing non-pharmaceutical interventions are marked with color.

**Figure S1:**
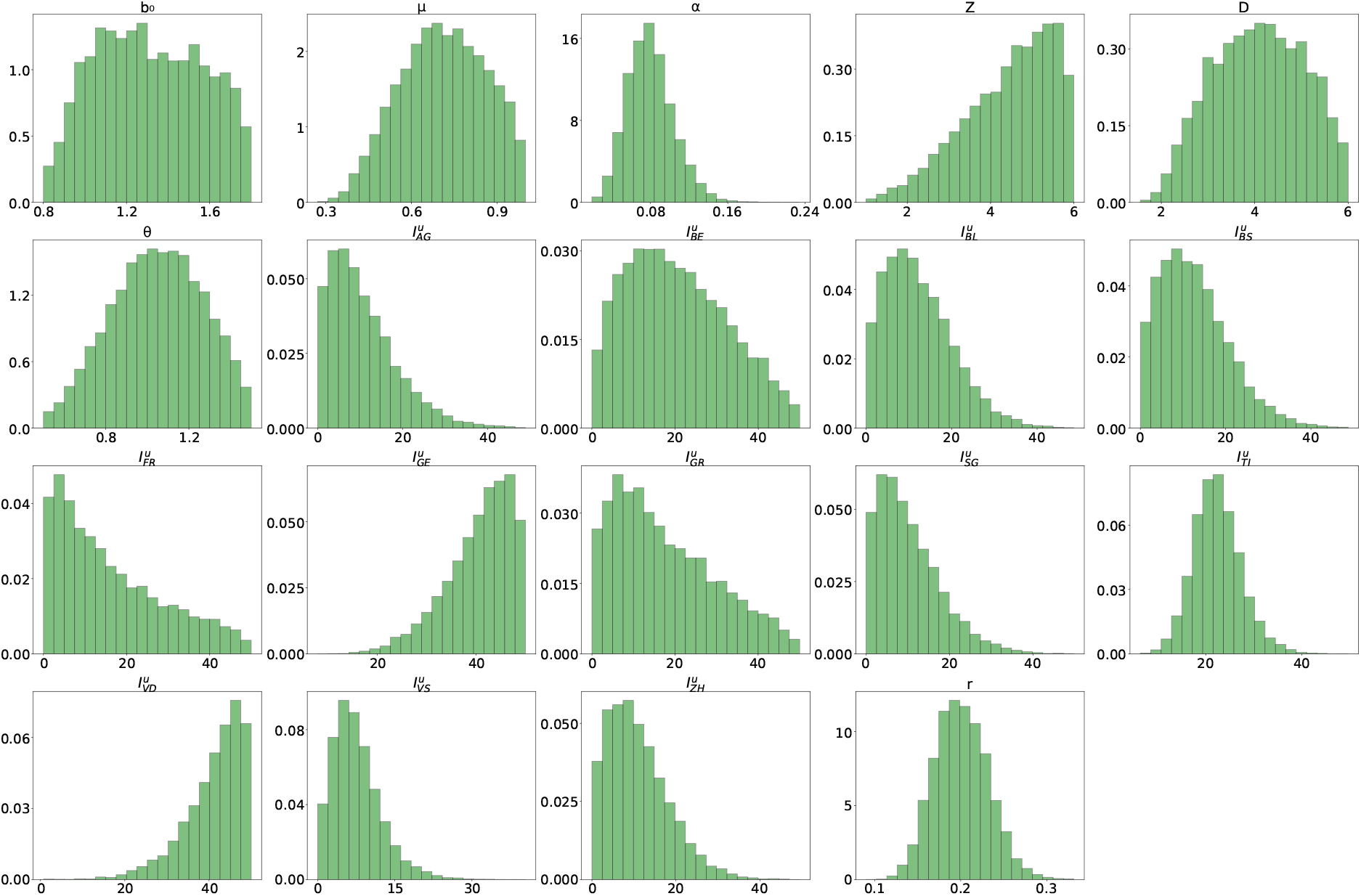
Marginal Figure S1: Marginal posterior distributions with data up to 17^th^ of March 2020. The used data correspond to the daily reported infectious persons in the cantons of Switzerland. The marginals with a canton label XY correspond to the initial condition 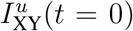 for the unreported cases in that canton.

**Figure S2:**
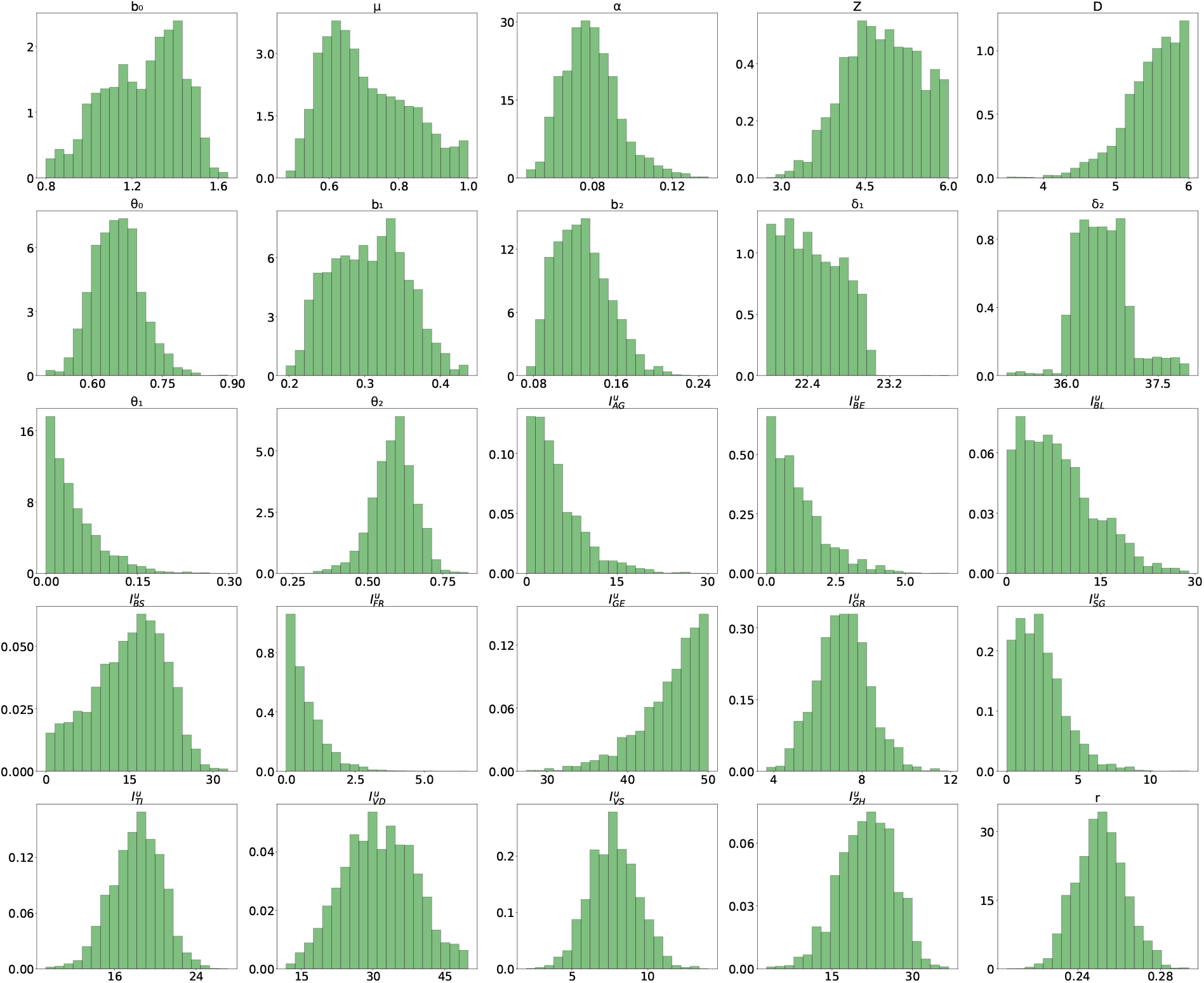
Marginal posterior distributions with data up to 6 ^th^ of June 2020. The used data correspond to the daily reported infectious persons in the cantons of Switzerland. The marginals with a canton label XY correspond to the initial condition 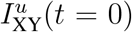 for the unreported cases in that canton.

**Figure S3:**
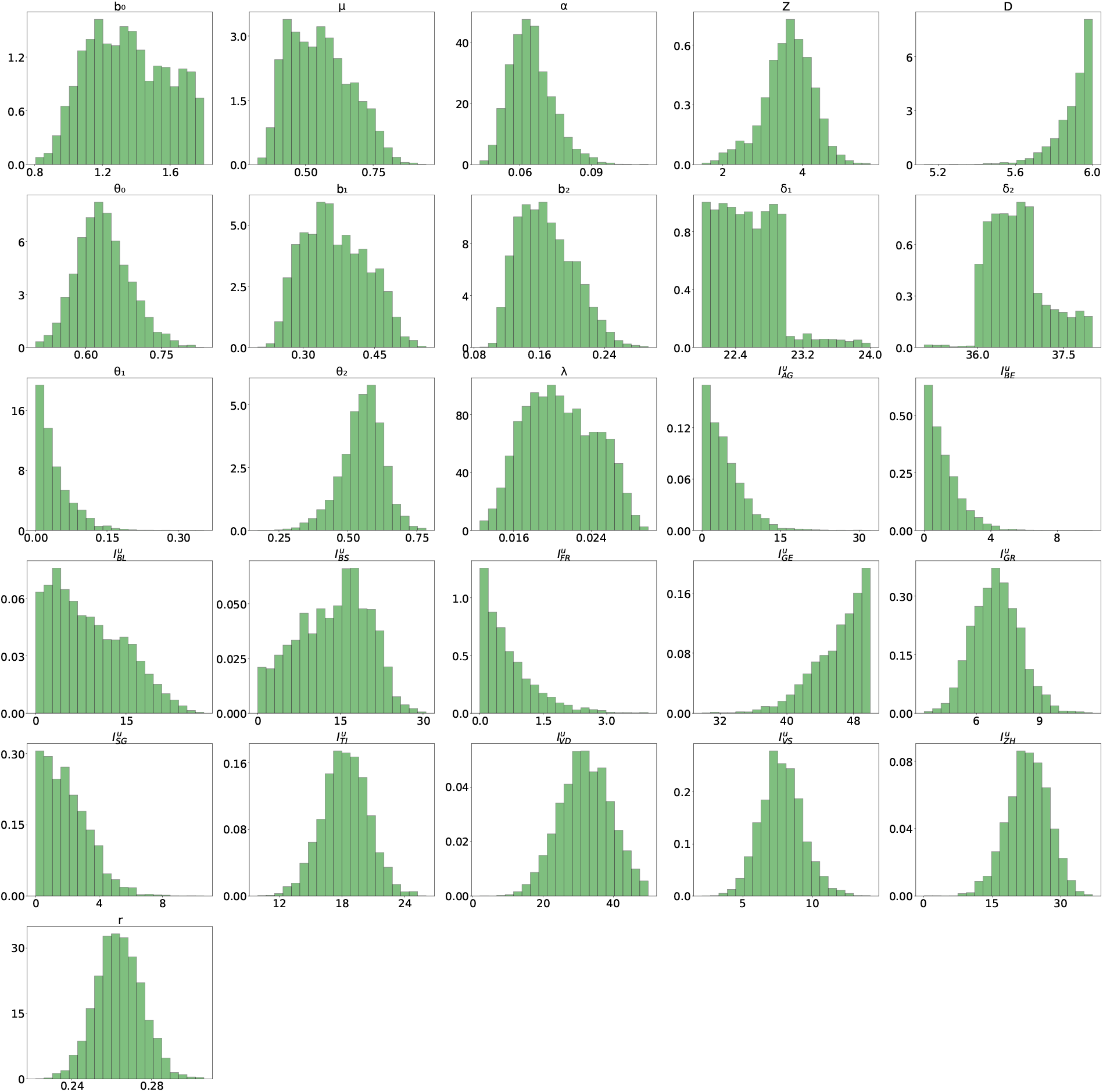
Marginal posterior distributions with data up to 9 ^th^ of July 2020. The used data correspond to the daily reported infectious persons in the cantons of Switzerland. The marginals with a canton label XY correspond to the initial condition 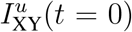 for the unreported cases in that canton.

**Figure S4:**
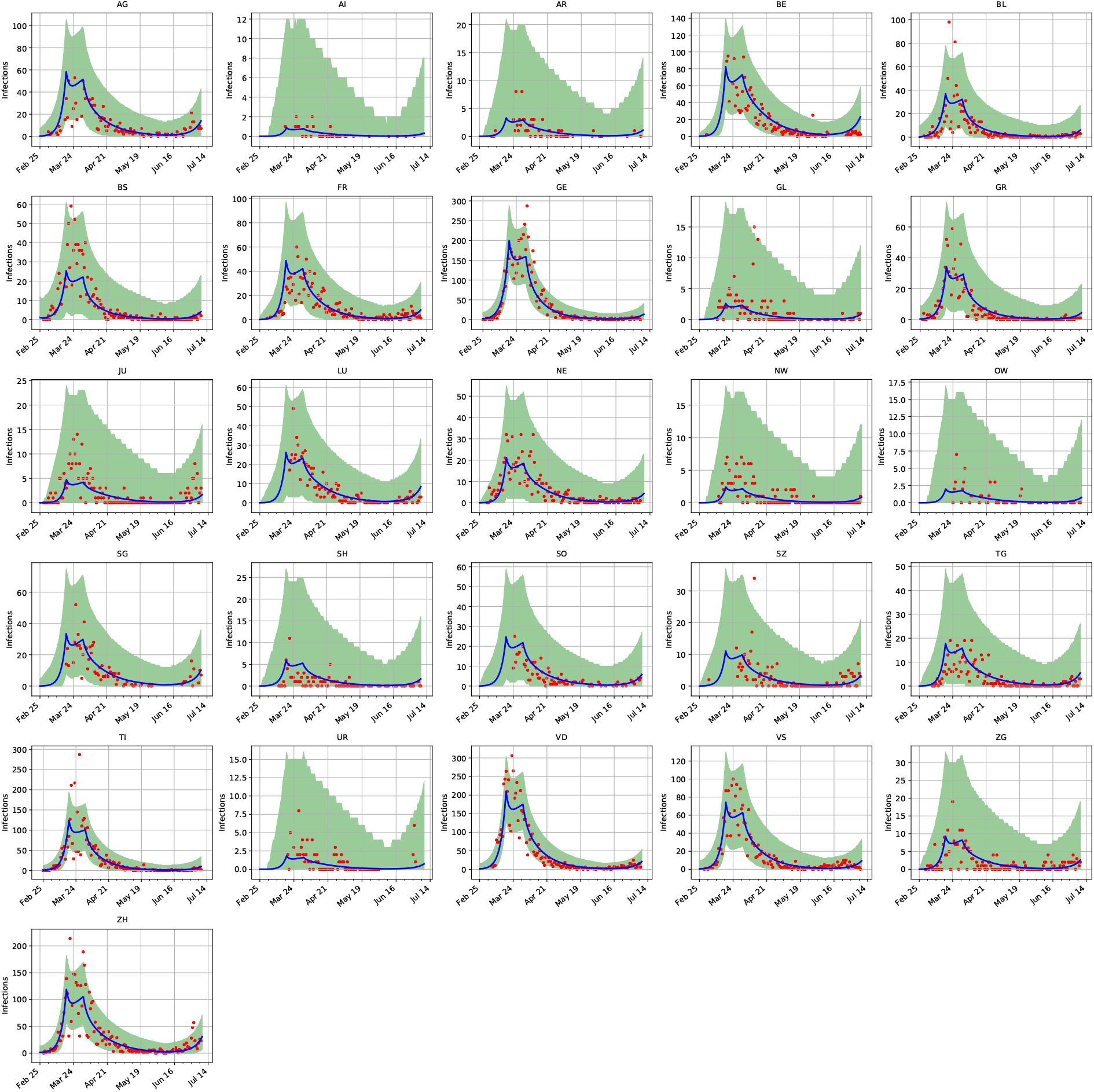
Maximum a-posteriori prediction with data up to 9^th^ of July 2020. The red points correspond to the daily reported cases per cantons and the blue curve shows the maximum aposteriori prediction. The 99% confidence interval is plotted in green and based on the sample shown in figure S3.

**Figure S5:**
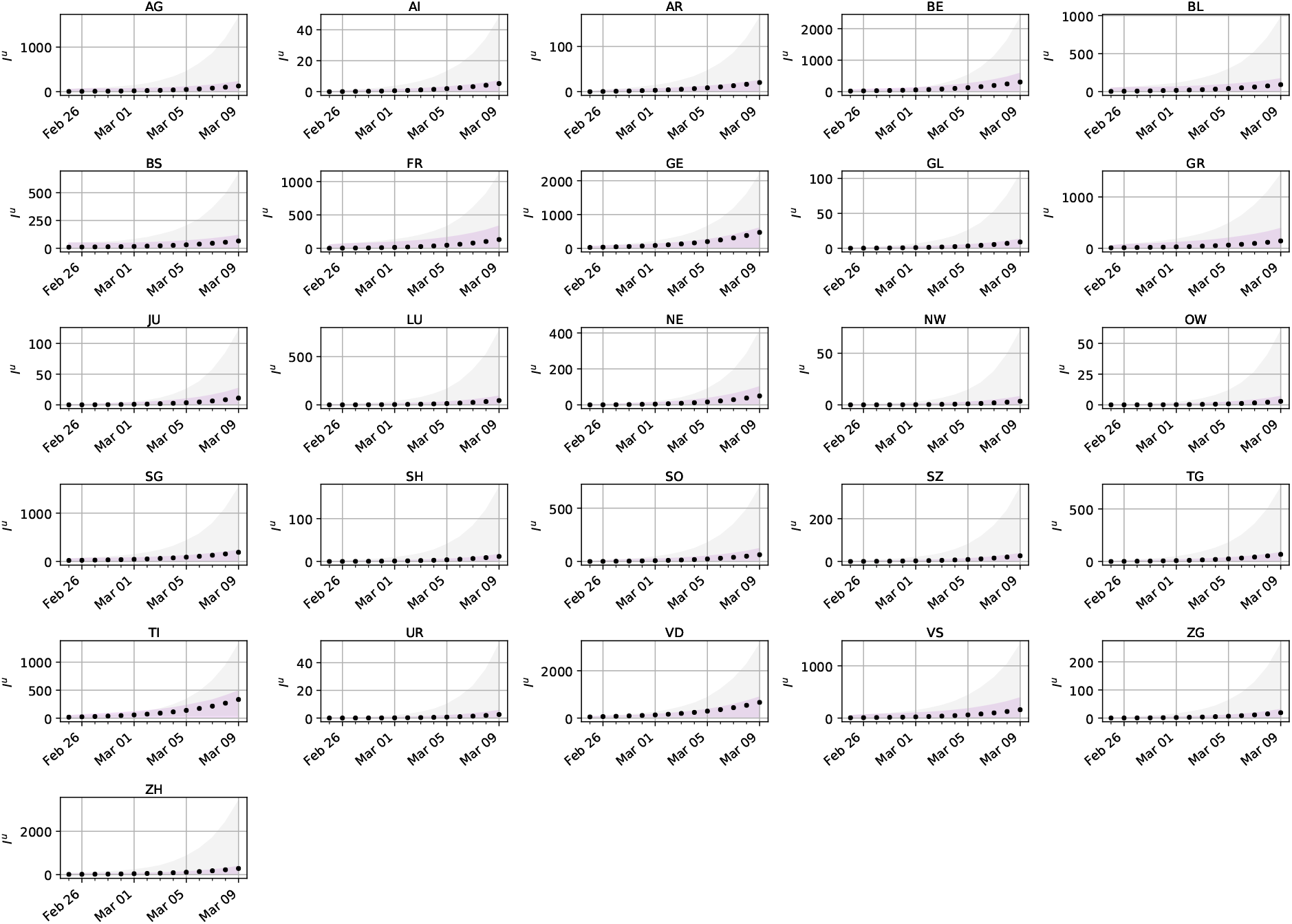
Comparison of prediction uncertainty per canton. The predictions are based on optimal strategies and non-specific testing for collection of data. They are also based on the *SEI*^*r*^*I*^*u*^*R* model output. The error bounds show the 99% confidence intervals of the unreported infectious model output for samples of the parameters with data obtained by optimal (purple) and standard testing (gray). The black dots show the actual unreported infectious for an artificial spread in Switzerland.

**Figure S6:**
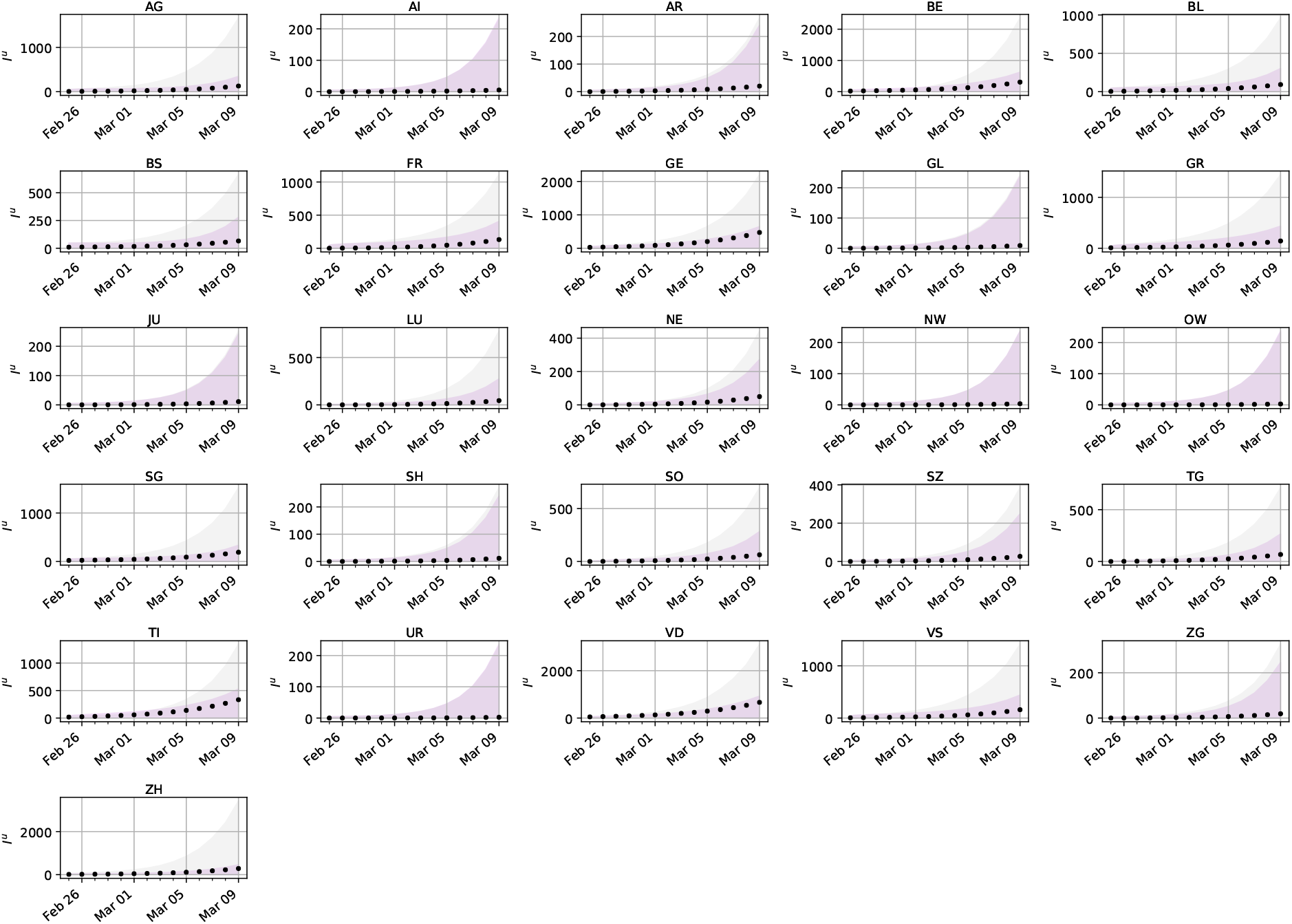
Comparison of propagated uncertainty per canton. The predictions are based on optimal strategies and non-specific testing. The *SEI*^*r*^*I*^*u*^*R* model output with added model error for the unreported infectious is shown. The error bounds show the 99% confidence intervals of the model output with added model error for samples of the parameters with data obtained by optimal (purple) and standard testing (gray). The black dots show the actual unreported infectious for an artificial spread in Switzerland.

**Table S1:**
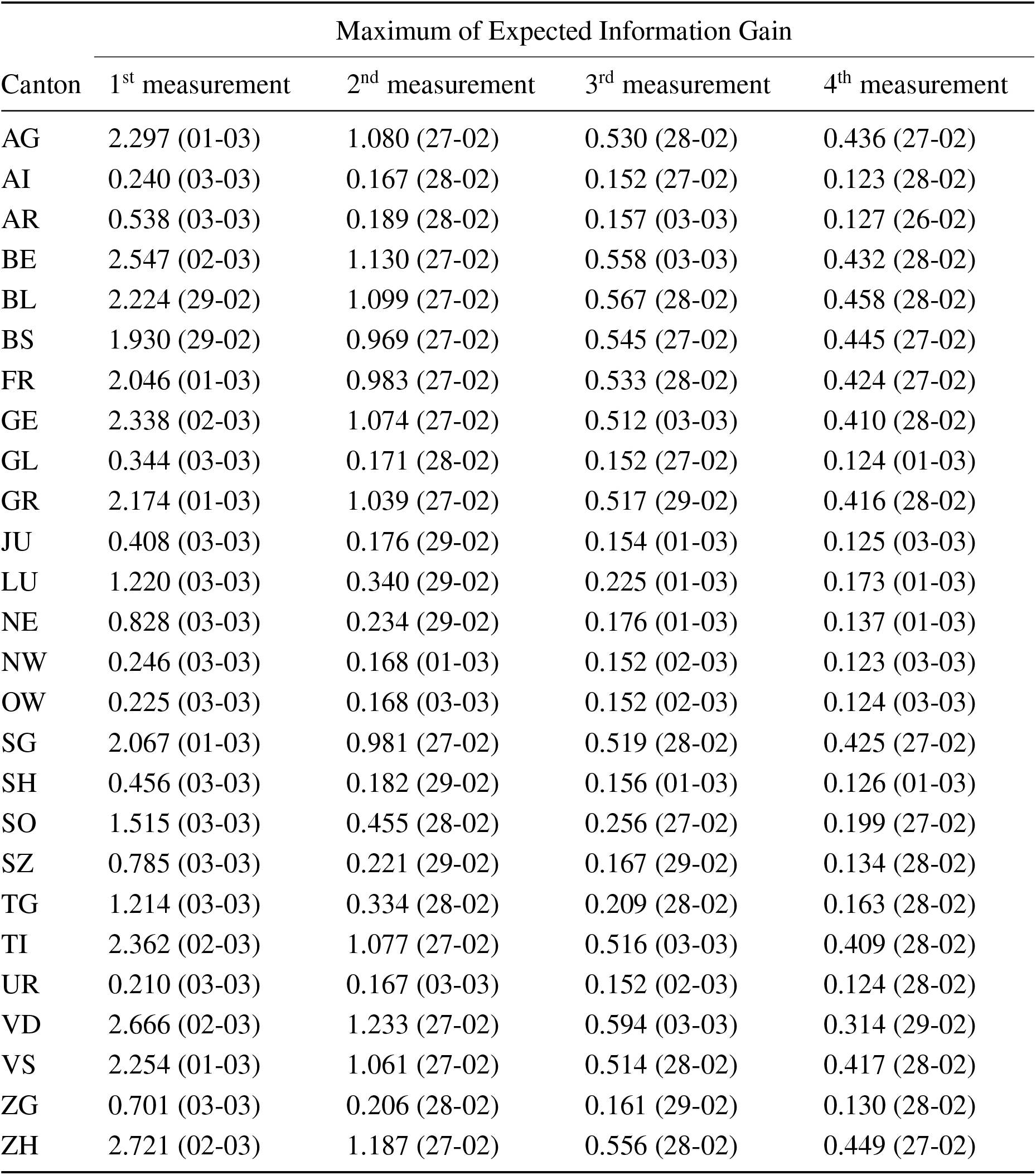
Maximum expected information gain for outbreak of a new disease. The shown expected information gain per measurement is defined in equation (17). The corresponding optimal dates are shown in parenthesis.

**Table S2:** Maximum expected information gain of non-pharmaceutical interventions. The corresponding optimal dates are shown in parenthesis.

| Canton | Maximum of Expected Information Gain |  |  |  |
| --- | --- | --- | --- | --- |
|  | 1<sup>st</sup> measurement | 2<sup>nd</sup> measurement | 3<sup>rd</sup> measurement | 4<sup>th</sup> measurement |
| AG | 2.307 (30-03) | 1.128 (17-03) | 0.399 (30-03) | 0.264 (30-03) |
| AI | 0.211 (30-03) | 0.169 (17-03) | 0.146 (21-03) | 0.112 (26-03) |
| AR | 0.474 (30-03) | 0.205 (17-03) | 0.154 (29-03) | 0.115 (29-03) |
| BE | 2.927 (30-03) | 1.738 (17-03) | 0.691 (17-03) | 0.459 (17-03) |
| BL | 1.663 (30-03) | 0.710 (17-03) | 0.241 (17-03) | 0.167 (17-03) |
| BS | 1.359 (30-03) | 0.518 (17-03) | 0.196 (17-03) | 0.140 (17-03) |
| FR | 2.149 (30-03) | 1.093 (17-03) | 0.256 (17-03) | 0.176 (17-03) |
| GE | 2.825 (29-03) | 1.760 (17-03) | 0.483 (21-03) | 0.327 (21-03) |
| GL | 0.364 (30-03) | 0.183 (17-03) | 0.149 (28-03) | 0.114 (29-03) |
| GR | 1.783 (30-03) | 1.256 (17-03) | 0.754 (17-03) | 0.222 (21-03) |
| JU | 0.736 (30-03) | 0.251 (17-03) | 0.163 (30-03) | 0.121 (30-03) |
| LU | 1.830 (30-03) | 0.738 (17-03) | 0.316 (30-03) | 0.210 (30-03) |
| NE | 1.573 (30-03) | 0.626 (17-03) | 0.205 (17-03) | 0.145 (17-03) |
| NW | 0.369 (30-03) | 0.181 (17-03) | 0.149 (30-03) | 0.114 (29-03) |
| OW | 0.332 (30-03) | 0.176 (17-03) | 0.148 (30-03) | 0.114 (29-03) |
| SG | 2.056 (30-03) | 1.003 (17-03) | 0.471 (17-03) | 0.247 (17-03) |
| SH | 0.625 (30-03) | 0.215 (17-03) | 0.158 (29-03) | 0.119 (30-03) |
| SO | 1.642 (30-03) | 0.663 (17-03) | 0.224 (30-03) | 0.155 (30-03) |
| SZ | 1.066 (30-03) | 0.330 (17-03) | 0.186 (30-03) | 0.135 (30-03) |
| TG | 1.495 (30-03) | 0.548 (17-03) | 0.250 (30-03) | 0.168 (30-03) |
| TI | 2.640 (29-03) | 1.639 (17-03) | 0.668 (17-03) | 0.436 (17-03) |
| UR | 0.315 (30-03) | 0.175 (17-03) | 0.148 (29-03) | 0.114 (29-03) |
| VD | 3.139 (30-03) | 1.961 (17-03) | 0.415 (22-03) | 0.311 (23-03) |
| VS | 2.313 (30-03) | 1.251 (17-03) | 0.397 (17-03) | 0.264 (17-03) |
| ZG | 0.920 (30-03) | 0.285 (17-03) | 0.172 (30-03) | 0.127 (30-03) |
| ZH | 2.980 (30-03) | 1.695 (17-03) | 0.735 (17-03) | 0.494 (17-03) |

**Table S3:**
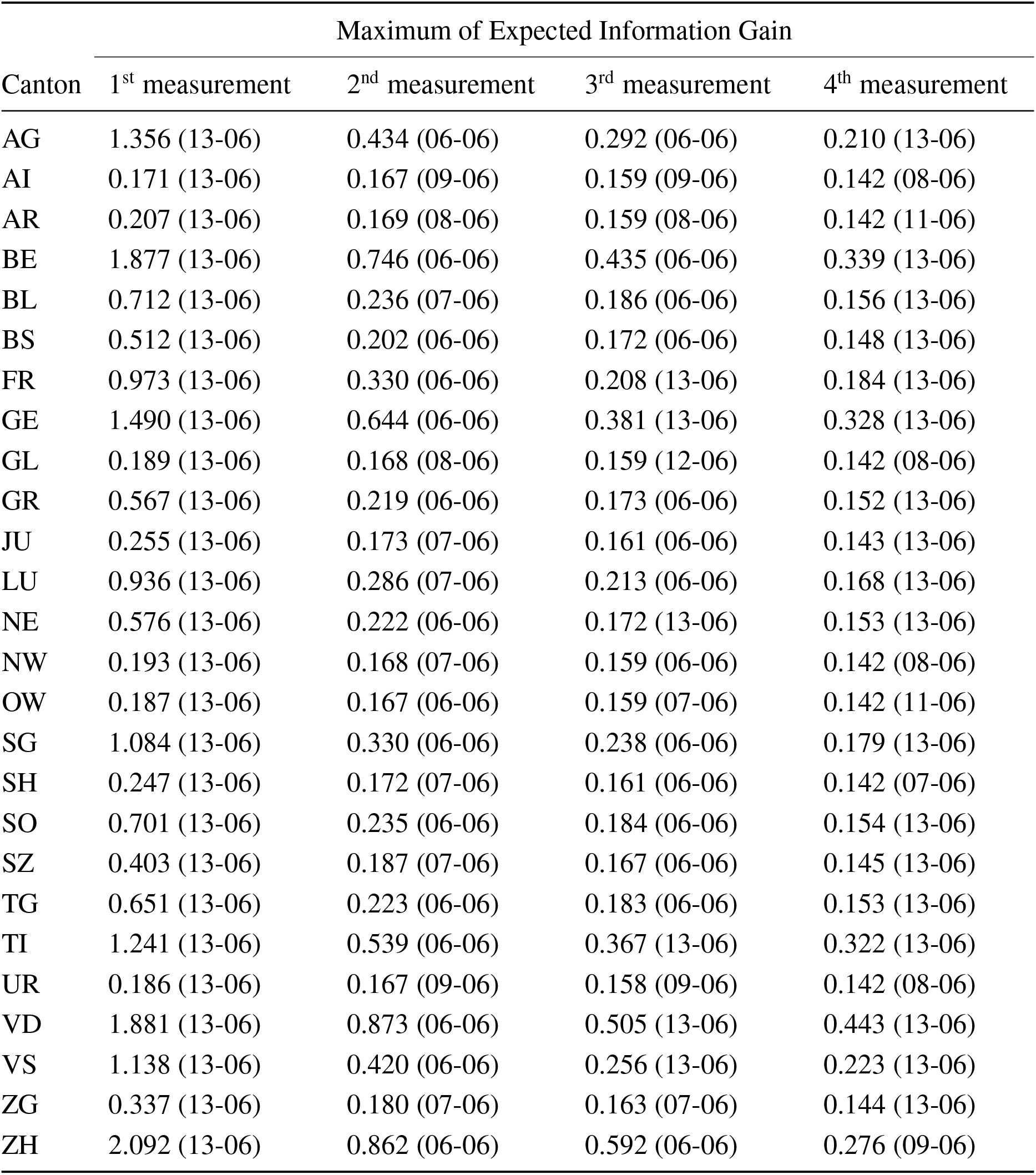
Maximum expected information gain for monitoring of a second outbreak with uninformed. *b*_3_. The corresponding optimal dates are shown in parenthesis.

**Table S4:**
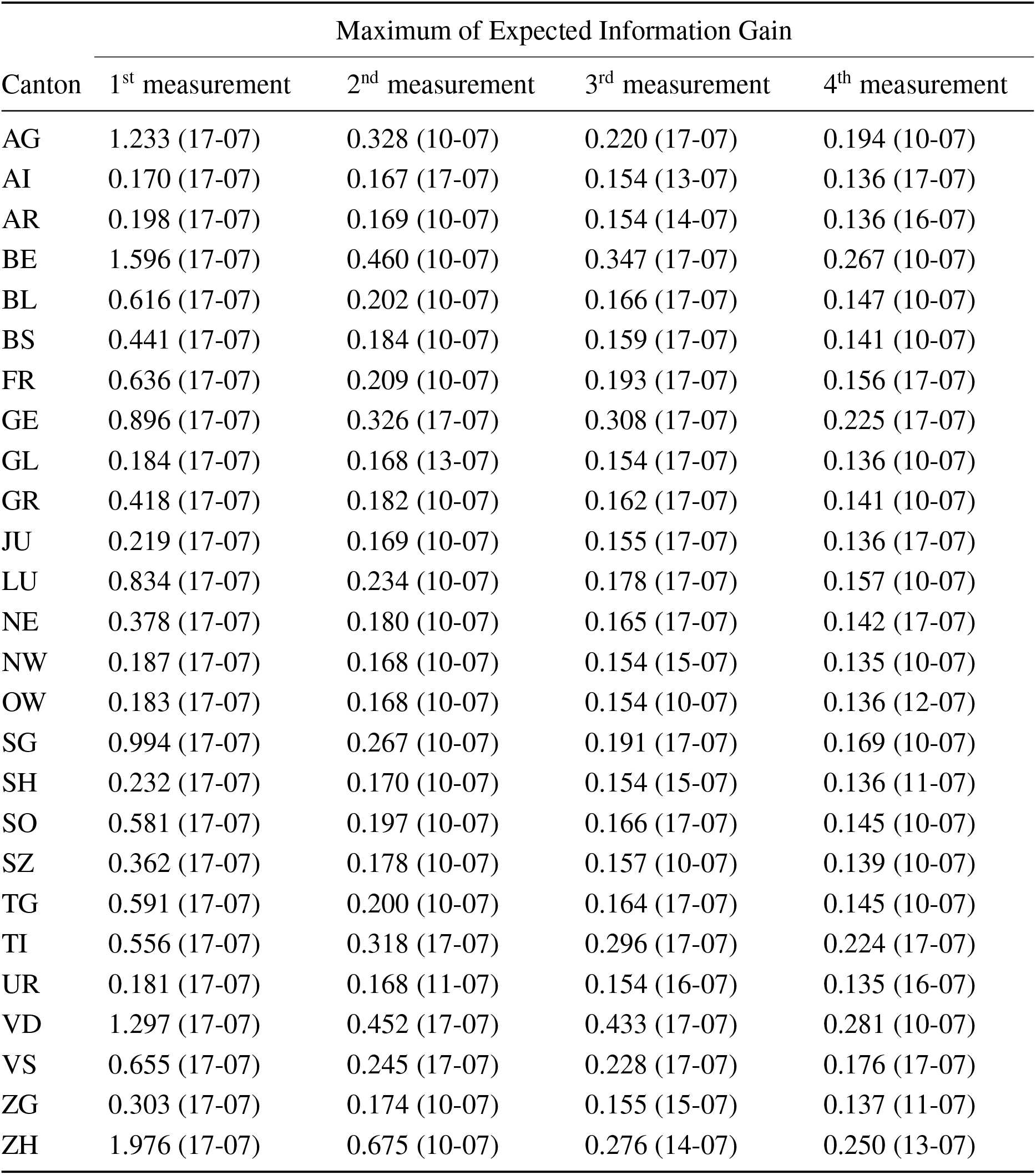
Maximum expected information gain to monitor a second outbreak with informed. *b*_3_. The corresponding optimal dates are shown in parenthesis.

| Canton | Maximum of Expected Information Gain |  |  |  |
| --- | --- | --- | --- | --- |
|  | 1<sup>st</sup> measurement | 2<sup>nd</sup> measurement | 3<sup>rd</sup> measurement | 4<sup>th</sup> measurement |
| AG | 1.233 (17-07) | 0.328 (10-07) | 0.220 (17-07) | 0.194 (10-07) |
| AI | 0.170 (17-07) | 0.167 (17-07) | 0.154 (13-07) | 0.136 (17-07) |
| AR | 0.198 (17-07) | 0.169 (10-07) | 0.154 (14-07) | 0.136 (16-07) |
| BE | 1.596 (17-07) | 0.460 (10-07) | 0.347 (17-07) | 0.267 (10-07) |
| BL | 0.616 (17-07) | 0.202 (10-07) | 0.166 (17-07) | 0.147 (10-07) |
| BS | 0.441 (17-07) | 0.184 (10-07) | 0.159 (17-07) | 0.141 (10-07) |
| FR | 0.636 (17-07) | 0.209 (10-07) | 0.193 (17-07) | 0.156 (17-07) |
| GE | 0.896 (17-07) | 0.326 (17-07) | 0.308 (17-07) | 0.225 (17-07) |
| GL | 0.184 (17-07) | 0.168 (13-07) | 0.154 (17-07) | 0.136 (10-07) |
| GR | 0.418 (17-07) | 0.182 (10-07) | 0.162 (17-07) | 0.141 (10-07) |
| JU | 0.219 (17-07) | 0.169 (10-07) | 0.155 (17-07) | 0.136 (17-07) |
| LU | 0.834 (17-07) | 0.234 (10-07) | 0.178 (17-07) | 0.157 (10-07) |
| NE | 0.378 (17-07) | 0.180 (10-07) | 0.165 (17-07) | 0.142 (17-07) |
| NW | 0.187 (17-07) | 0.168 (10-07) | 0.154 (15-07) | 0.135 (10-07) |
| OW | 0.183 (17-07) | 0.168 (10-07) | 0.154 (10-07) | 0.136 (12-07) |
| SG | 0.994 (17-07) | 0.267 (10-07) | 0.191 (17-07) | 0.169 (10-07) |
| SH | 0.232 (17-07) | 0.170 (10-07) | 0.154 (15-07) | 0.136 (11-07) |
| SO | 0.581 (17-07) | 0.197 (10-07) | 0.166 (17-07) | 0.145 (10-07) |
| SZ | 0.362 (17-07) | 0.178 (10-07) | 0.157 (10-07) | 0.139 (10-07) |
| TG | 0.591 (17-07) | 0.200 (10-07) | 0.164 (17-07) | 0.145 (10-07) |
| TI | 0.556 (17-07) | 0.318 (17-07) | 0.296 (17-07) | 0.224 (17-07) |
| UR | 0.181 (17-07) | 0.168 (11-07) | 0.154 (16-07) | 0.135 (16-07) |
| VD | 1.297 (17-07) | 0.452 (17-07) | 0.433 (17-07) | 0.281 (10-07) |
| VS | 0.655 (17-07) | 0.245 (17-07) | 0.228 (17-07) | 0.176 (17-07) |
| ZG | 0.303 (17-07) | 0.174 (10-07) | 0.155 (15-07) | 0.137 (11-07) |
| ZH | 1.976 (17-07) | 0.675 (10-07) | 0.276 (14-07) | 0.250 (13-07) |

**Table S5:** Parameters and prior distributions used in Bayesian inference. Here the data corresponds to the daily reported infections. In all cases, data are used from the 25^th^ of February 2020, when the first reported case was found in the canton of Ticino. Inference I uses data up to the day non-pharmaceutical interventions were announced (17^th^ of March 2020). Inference II uses data up to the day measures were relaxed (6^th^ of June 2020). Inference III uses data up to the 9^th^ of July 2020. The choice of prior distributions is consistent with the choice found in (*14*); the ranges used in our study are slightly extended.

| Parameter | Prior distribution / fixed value | Inference I | Inference II | Inference III |
| --- | --- | --- | --- | --- |
| $b_0$ | $\mathcal{U}([0.8, 1.8])$ | yes | yes | yes |
| $\alpha$ | $\mathcal{U}([0.01, 1])$ | yes | yes | yes |
| $\mu$ | $\mathcal{U}([0.2, 1])$ | yes | yes | yes |
| $Z$ | $\mathcal{U}([1, 6])$ | yes | yes | yes |
| $D$ | $\mathcal{U}([1, 6])$ | yes | yes | yes |
| $\theta_0$ | $\mathcal{U}([0.5, 1.5])$ | yes | yes | yes |
| $c_1$ | $\mathcal{U}([0, 1])$ | no | yes | yes |
| $c_2$ | $\mathcal{U}([0, 1])$ | no | yes | yes |
| $c_3$ | $\mathcal{U}([0, 1])$ | no | yes | yes |
| $c_4$ | $\mathcal{U}([0, 1])$ | no | yes | yes |
| $\delta_1$ | $\mathcal{U}([20, 30])$ | no | yes | yes |
| $\delta_2$ | $\mathcal{U}([30, 40])$ | no | yes | yes |
| $\delta_3$ | 102 | no | yes | yes |
| $\lambda$ | $\mathcal{U}([0, 0.03])$ | no | no | yes |
| $r$ | $\mathcal{U}([0, 2])$ | yes | yes | yes |
| $I_{IC}^u$ | $\mathcal{U}([0, 50]^{12})$ | yes | yes | yes |

